# Early Lymphocyte Dynamics and Subsequent Cancer Therapy–Related Cardiac Dysfunction in Patients Receiving Anthracycline Therapy

**DOI:** 10.64898/2026.10.26.26366568

**Authors:** Shilan Zhang, Ling Dao, Yanan Pang, Sensen Zhang, Bing Shi, Jingwen Li, Lei Hou

## Abstract

**Background:** Whether early treatment-related lymphocyte dynamics are associated with cancer therapy–related cardiac dysfunction (CTRCD) after anthracycline therapy is uncertain. This study sought to determine the association between early change in lymphocyte and subsequent CTRCD and to assess its incremental value beyond clinical characteristics.

**Methods:** We retrospectively studied anthracycline-treated adults in the Medical Information Mart for Intensive Care IV (MIMIC-IV) database with linked echocardiographic data (MIMIC-IV-Echo). The primary exposure was relative change in lymphocyte percentage from baseline to the 0–30-day nadir. CTRCD was defined as a left ventricular ejection fraction (LVEF) decline ≥10 percentage points to <53%. Associations were modeled using multivariable logistic regression with restricted cubic splines. Incremental discrimination was assessed using optimism-corrected AUCs.

**Results:** Among 362 patients in the analysis cohort, 58 (16.0%) developed CTRCD. Baseline lymphocyte measures were not associated with CTRCD. Early lymphocyte percentage change was associated with CTRCD (overall P=0.031). Compared with the median change (−73%), a less pronounced decline at the 90th percentile (−5%) was associated with higher odds of CTRCD (OR: 1.96; 95% CI: 1.12–3.42; P=0.018). The association remained consistent after sequential and hematologic adjustment. Added to a clinical model that included hemoglobin and immature granulocyte percentage, lymphocyte change increased the apparent AUC from 0.705 to 0.724 and the optimism-corrected AUC from 0.644 to 0.658 (P = 0.040).

**Conclusions:** A less pronounced early decline in lymphocyte percentage after anthracycline initiation was associated with higher odds of subsequent CTRCD and provided modest incremental information beyond clinical characteristics.

**Clinical Perspective:** *What Is New?:* A less pronounced early decline in lymphocyte percentage during the first 30 days after anthracycline initiation was associated with higher odds of subsequent cancer therapy–related cardiac dysfunction (CTRCD).

*What Are the Clinical Implications?:* Early lymphocyte dynamics may provide modest incremental information beyond clinical characteristics, but prospective validation is needed before clinical application.

**Graphic Abstract. Early Lymphocyte Response and Subsequent CTRCD After Anthracycline Therapy.:** Among 362 adults with a recorded inpatient anthracycline order in MIMIC-IV, the primary exposure was the relative change in lymphocyte percentage from baseline to the nadir during days 0–30. CTRCD was defined as an LVEF decline of at least 10 percentage points to below 53%, assessed during days 31–365. A less pronounced early decline in lymphocyte percentage was associated with higher odds of subsequent CTRCD (overall association P = 0.031; OR, 1.96; 95% CI, 1.12–3.42, comparing the 90th with the 50th percentile). Adding lymphocyte change to the clinical model, which included baseline hemoglobin and immature granulocyte percentage, increased the optimism-corrected AUC from 0.644 to 0.658.

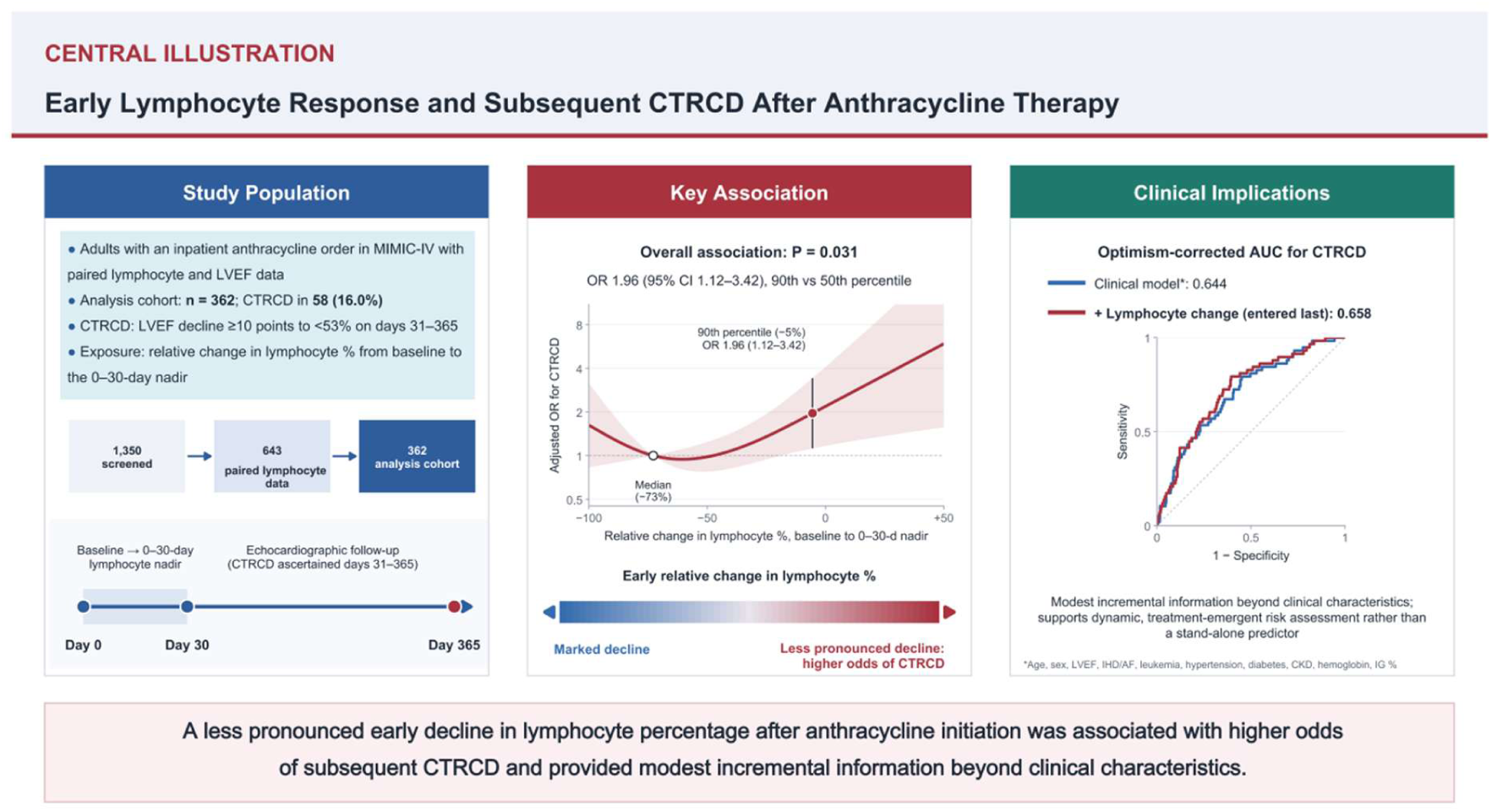

## 1 Introduction

Anthracyclines remain a cornerstone of therapy for a broad range of hematologic and solid malignancies, but their clinical use is limited by cancer therapy–related cardiac dysfunction (CTRCD). Contemporary cardio-oncology practice emphasizes baseline cardiovascular risk assessment and serial surveillance using cardiac imaging and biomarkers during anthracycline therapy (1–3). However, susceptibility to cardiotoxicity evolves over the treatment course, and recent evidence suggests that post-treatment reassessment can provide additional information beyond baseline risk assessment (1,4).

Growing evidence indicates that anthracycline-related cardiac injury involves not only direct cardiomyocyte toxicity but also innate and adaptive immune activation. Recent mechanistic work has implicated neutrophils, monocytes, lymphocytes, and other immune-cell populations in the inflammatory and remodeling responses to anthracycline exposure (5). Peripheral blood-cell profiles may therefore provide a readily available window into treatment-associated systemic responses. In patients with breast cancer, pretreatment lymphocyte-derived inflammatory indices have been associated with CTRCD (6), whereas our previous study found that post-treatment lymphocyte-based immunoinflammatory indices showed greater discriminatory value for CTRCD than their pretreatment counterparts (7).

These prior approaches have generally evaluated lymphocyte-related markers at discrete pretreatment or post-treatment time points or as derived inflammatory ratios. Such approaches characterize hematologic status but do not directly quantify the magnitude of an individual patient’s early response after treatment initiation. Accordingly, we investigated the association between early within-patient relative change in lymphocyte percentage during the first 30 days after anthracycline initiation and subsequent CTRCD. We further assessed whether this dynamic measure provided incremental discrimination beyond clinical characteristics.

## 2 Methods

### 2.1 Data Source and Study Population

This retrospective cohort study used deidentified electronic health record data from the MIMIC-IV database linked to structured echocardiographic measurements from MIMIC-IV-Echo (8). The MIMIC-IV data resource was approved by the institutional review boards of Beth Israel Deaconess Medical Center and the Massachusetts Institute of Technology, with a waiver of individual informed consent because the data were deidentified.

Adults aged ≥18 years with a recorded inpatient order for doxorubicin, daunorubicin, epirubicin, idarubicin, or mitoxantrone were eligible for inclusion. The timestamp of the first qualifying anthracycline order was defined as the index date. Patients with pre-existing heart failure recorded on or before the index date were excluded. The analysis cohort additionally required an available baseline lymphocyte percentage, at least 1 lymphocyte percentage measurement during the first 30 days after the index date, and paired baseline and follow-up LVEF measurements for CTRCD ascertainment.

### 2.2 Outcome and Primary Exposure Definitions

The primary outcome was CTRCD, operationally defined as a decline in LVEF of ≥10 percentage points from baseline to an absolute value < 53% on follow-up echocardiography performed between days 31 and 365 after anthracycline initiation, based on the ASE/EACVI LVEF-based criteria (9). An LVEF-based definition was used because global longitudinal strain and cardiac biomarkers were not sufficiently available to apply a contemporary multiparametric CTRCD definition. For patients with multiple qualifying follow-up examinations, the first echocardiogram meeting the CTRCD definition was used to determine CTRCD ascertainment. Baseline LVEF was defined as the measurement closest to the index date between 180 days before and 2 days after the index date. The primary exposure was the early within-patient relative change in lymphocyte percentage. Baseline lymphocyte percentage was defined as the measurement closest to the index date between 30 days before and 2 days after the index date, and the early post-treatment value was defined as the lowest lymphocyte percentage recorded during days 0 to 30 after anthracycline initiation. Relative change was calculated as 100 × (nadir − baseline) / baseline. Because the exposure window (days 0 to 30) ended before outcome ascertainment began (day 31), this design corresponds to a 30-day landmark analysis and avoids immortal-time bias.

### 2.3 Clinical and Hematologic Covariates

The primary model adjusted for age, sex, baseline LVEF, prior ischemic heart disease or atrial fibrillation, leukemia, hypertension, diabetes mellitus, and chronic kidney disease. For incremental discrimination analyses, the clinical model comprised these covariates plus baseline hemoglobin and immature granulocyte percentage. Baseline hemoglobin and immature granulocyte percentage were also examined as additional covariates in sensitivity analyses of the association. Availability of observed model covariates is summarized in Supplemental Table S6.

### 2.4 Statistical Analysis

Continuous variables are presented as median (Q1–Q3) and categorical variables as number (percentage), with between-group comparisons using the Wilcoxon rank-sum test or Fisher exact test, as appropriate. The association between early lymphocyte relative change and CTRCD was evaluated using multivariable logistic regression adjusted for the clinical covariates listed above. Lymphocyte change was winsorized at the 1st and 99th percentiles and modeled with restricted cubic splines using knots at the 10th, 50th, and 90th percentiles. Effect estimates are reported as ORs comparing the 90th with the 50th percentile of relative change. All covariates in the primary model were complete. In sensitivity analyses that additionally adjusted for baseline immature granulocyte percentage (missing in 110 patients [30.4%]), missing values were multiply imputed across 20 datasets by predictive mean matching and estimates were pooled using Rubin’s rules; complete-case analysis was also performed. Further sensitivity analyses added baseline hemoglobin and relative changes in other hematopoietic lineages, adjusted for cumulative glucocorticoid dose (prednisone equivalents) and granulocyte colony-stimulating factor orders during days 0 to 30, and applied inverse probability of selection weighting based on baseline characteristics and lymphocyte measures to account for the availability of paired LVEF measurements. Additional models adjusted for covariates not included in the primary model, including race, recorded inpatient anthracycline administration days and doxorubicin-equivalent dose, chronic obstructive pulmonary disease, red cell distribution width, smoking, obesity, valvular heart disease, prior myocardial infarction, and the number of days with lymphocyte measurements. Exploratory subgroup analyses compared patients with relatively preserved lymphocytes (relative change at or above the 75th percentile) with other patients, with effect modification assessed by likelihood ratio tests. Robustness to unmeasured confounding was quantified with the E-value. Incremental discrimination was assessed by adding the lymphocyte spline terms to the clinical model. Clinical covariates were also entered stepwise (demographics, cardiac status, malignancy and comorbidities, and baseline hematologic measures), with lymphocyte change entered last. Because baseline immature granulocyte percentage was part of the clinical model, missing values were multiply imputed as described above and predicted probabilities were averaged across the 20 imputed datasets. AUCs were corrected for optimism by bootstrap (250 resamples in each imputed dataset; 5,000 in total), the contribution of the lymphocyte spline terms was tested with a pooled Wald test, continuous net reclassification improvement (NRI) and integrated discrimination improvement (IDI) were estimated with 95% CIs from 500 bootstrap resamples, and the calibration slope was bootstrap-corrected. All tests were 2-sided, with P < 0.05 considered statistically significant. Analyses were performed using R version 4.5.2.

## 3 Results

### 3.1 Study Cohort and Baseline Characteristics

Of 1,350 patients with a recorded inpatient anthracycline order, 143 with pre-existing heart failure and 564 without paired lymphocyte measurements were excluded. Among the remaining 643 patients, 279 lacked paired baseline and follow-up LVEF measurements and 2 had an undefined relative lymphocyte change (baseline and nadir values both 0%), yielding an analysis cohort of 362 patients. Of these, 58 (16.0%) met the definition of CTRCD and 304 (84.0%) did not (Figure 1).

**Figure 1.**
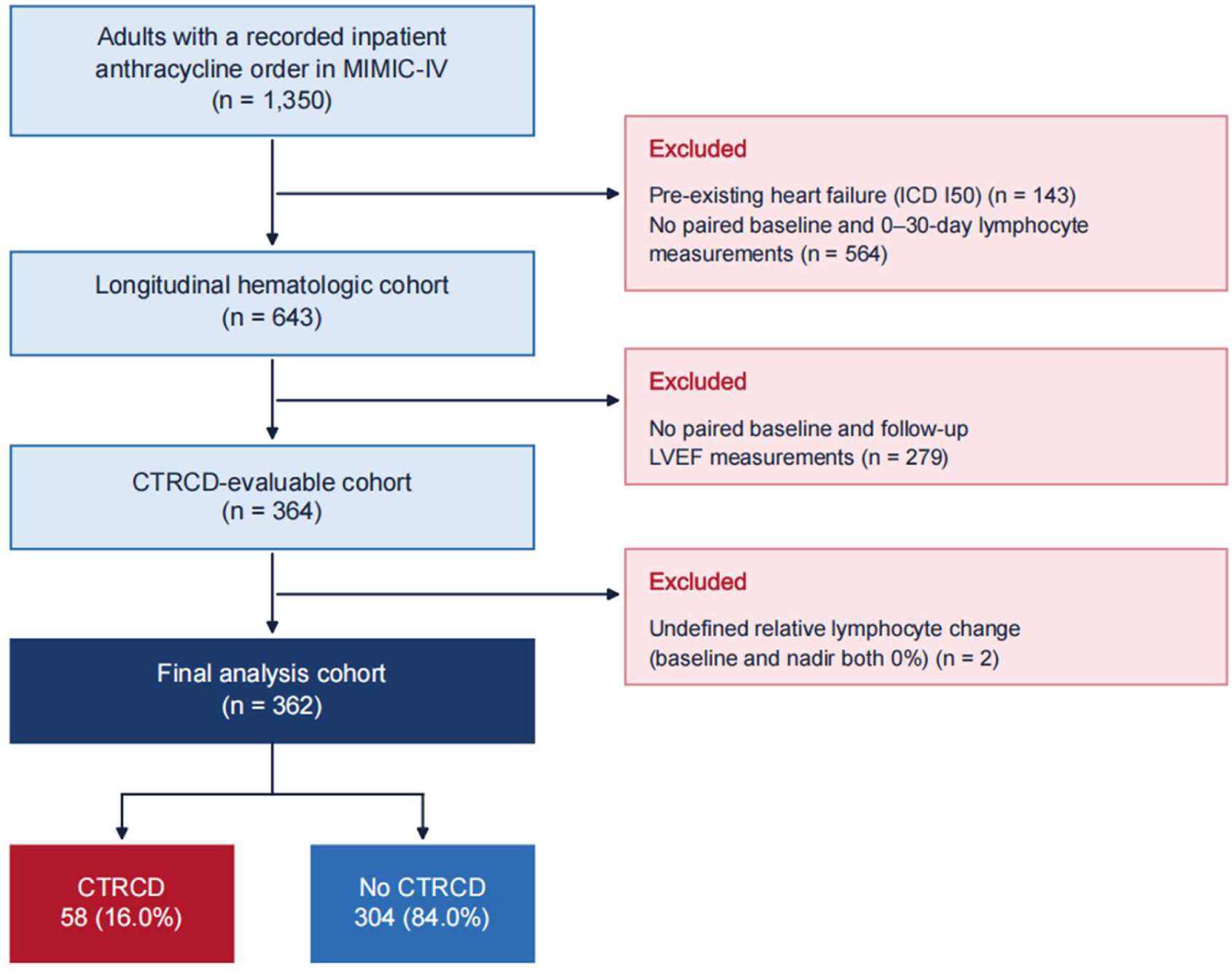
Cohort Derivation. Flow diagram showing the derivation of the analysis cohort from 1,350 adults with a recorded inpatient anthracycline order in MIMIC-IV. After excluding 143 patients with pre-existing heart failure and 564 without paired baseline and 0–30-day lymphocyte measurements, 643 patients remained. A further 279 patients without paired baseline and follow-up left ventricular ejection fraction (LVEF) measurements and 2 with undefined relative lymphocyte change were excluded. The final analysis cohort comprised 362 patients, including 58 (16.0%) with cancer therapy–related cardiac dysfunction (CTRCD) and 304 (84.0%) without CTRCD. MIMIC-IV = Medical Information Mart for Intensive Care IV.

The median age was 61 years (Q1–Q3: 50–69 years), 175 patients (48.3%) were female, and lymphoma (56.6%) and leukemia (32.9%) were the predominant malignancies. Baseline lymphocyte measures and lymphocyte-derived inflammatory indices did not differ by subsequent CTRCD status. Patients who developed CTRCD more frequently had ischemic heart disease (13.8% vs 4.9%; P = 0.018) and had lower hemoglobin (8.4 [7.6–9.6] vs 9.3 [8.0–11.1] g/dL; P = 0.001) and higher immature granulocyte percentage (1.1% [0.7–1.9] vs 0.7% [0.5–1.3]; P = 0.010). They were also less often White (58.6% vs 75.0%; P = 0.016), more frequently had chronic obstructive pulmonary disease (8.6% vs 2.6%; P = 0.041), had higher red cell distribution width (16.6% vs 14.9%; P = 0.017), and had fewer recorded inpatient anthracycline administration days (median, 1 vs 2; P < 0.001). Baseline LVEF and recorded inpatient doxorubicin-equivalent dose were similar between groups (Table 1).

**Table 1.** Baseline Characteristics According to CTRCD Status.

| Characteristic | Overall<br>N = 362 | No CTRCD<br>N = 304 | CTRCD<br>N = 58 | P value |
| --- | --- | --- | --- | --- |
| **Demographics** |  |  |  |  |
| Age at anthracycline initiation, y | 61 (50–69) | 61 (50–69) | 61 (50–70) | 0.856 |
| **Sex** |  |  |  | **0.570** |
| Female | 175 (48.3) | 149 (49.0) | 26 (44.8) |  |
| Male | 187 (51.7) | 155 (51.0) | 32 (55.2) |  |
| **Race** |  |  |  | 0.016 |
| White | 262 (72.4) | 228 (75.0) | 34 (58.6) |  |
| Non-White or unknown | 100 (27.6) | 76 (25.0) | 24 (41.4) |  |
| **Smoking** |  |  |  |  |
| Tobacco use | 33 (9.1) | 26 (8.6) | 7 (12.1) | 0.453 |
| No tobacco use | 329 (90.9) | 278 (91.4) | 51 (87.9) |  |
| **Drinking** |  |  |  |  |
| Alcohol use | 16 (4.4) | 14 (4.6) | 2 (3.4) | >0.99 |
| No alcohol use | 346 (95.6) | 290 (95.4) | 56 (96.6) |  |
| **Insurance (socioeconomic proxy)** |  |  |  |  |
| Medicaid | 61 (16.9) | 43 (14.1) | 18 (31.0) | 0.003 |
| Medicare, private, or other | 301 (83.1) | 261 (85.9) | 40 (69.0) |  |
| **Underlying malignancy** |  |  |  |  |
| Lymphoma | 205 (56.6) | 178 (58.6) | 27 (46.6) | 0.112 |
| Leukemia | 119 (32.9) | 94 (30.9) | 25 (43.1) | 0.093 |
| Multiple myeloma | 9 (2.5) | 7 (2.3) | 2 (3.4) | 0.641 |
| Sarcoma | 15 (4.1) | 14 (4.6) | 1 (1.7) | 0.482 |
| Breast cancer | 5 (1.4) | 3 (1.0) | 2 (3.4) | 0.183 |
| Other or unspecified | 25 (6.9) | 22 (7.2) | 3 (5.2) | 0.779 |
| **Comorbidities** |  |  |  |  |
| Hypertension | 155 (42.8) | 130 (42.8) | 25 (43.1) | >0.99 |
| Diabetes mellitus | 64 (17.7) | 54 (17.8) | 10 (17.2) | >0.99 |
| Chronic kidney disease | 29 (8.0) | 23 (7.6) | 6 (10.3) | 0.436 |
| Ischemic heart disease | 23 (6.4) | 15 (4.9) | 8 (13.8) | 0.018 |
| Atrial fibrillation | 39 (10.8) | 31 (10.2) | 8 (13.8) | 0.486 |
| Obesity | 44 (12.2) | 40 (13.2) | 4 (6.9) | 0.271 |
| Prior myocardial infarction | 13 (3.6) | 10 (3.3) | 3 (5.2) | 0.446 |
| Valvular heart disease | 24 (6.6) | 22 (7.2) | 2 (3.4) | 0.395 |
| COPD | 13 (3.6) | 8 (2.6) | 5 (8.6) | 0.041 |
| **Medication use** |  |  |  |  |
| ACEI, ARB, or ARNI | 51 (14.1) | 43 (14.1) | 8 (13.8) | >0.99 |
| β-Blocker | 82 (22.7) | 67 (22.0) | 15 (25.9) | 0.499 |
| Statin | 63 (17.4) | 53 (17.4) | 10 (17.2) | >0.99 |
| **Baseline cardiac assessment** |  |  |  |  |
| Baseline LVEF, % | 62 (59–68) | 62 (58–68) | 64 (60–66) | 0.953 |
| **Baseline Hematologic parameters** |  |  |  |  |
| Hemoglobin, g/dL | 9.1 (7.9–10.8) | 9.3 (8.0–11.1) | 8.4 (7.6–9.6) | 0.001 |
| Platelet count, ×10<sup>9</sup>/L | 180 (84–292) | 188 (90–306) | 162 (60–230) | 0.084 |
| White blood cell count, ×10<sup>9</sup>/L | 7.3 (4.3–11.2) | 7.2 (4.3–11.1) | 7.4 (4.1–12.6) | 0.789 |
| RDW, % | 15.0 (13.4– | 14.9 (13.4–17.0) | 16.6 (14.1–18.3) | 0.017 |
| ALC, ×10<sup>9</sup>/L | 0.74 (0.38– | 0.78 (0.39–1.37) | 0.60 (0.34–1.89) | 0.807 |
| Lymphocyte, % | 10.9 (5.4–22.6) | 11.6 (5.5–22.0) | 9.3 (5.0–24.0) | 0.436 |
| Neutrophil, % | 74.9 (53.0– | 76.1 (53.2–87.8) | 69.1 (31.0–88.7) | 0.473 |
| Monocyte, % | 5.0 (2.0–9.1) | 5.0 (2.2–9.1) | 5.0 (2.0–8.7) | 0.760 |
| Immature granulocyte, % | 0.7 (0.5–1.3) | 0.7 (0.5–1.3) | 1.1 (0.7–1.9) | 0.010 |
| NLR | 5.9 (1.8–14.5) | 5.9 (1.9–14.0) | 5.7 (1.3–15.6) | 0.787 |
| PLR | 237 (87–520) | 239 (99–517) | 235 (47–557) | 0.473 |
| LMR | 2.3 (1.2–5.0) | 2.2 (1.2–4.5) | 2.6 (1.3–5.5) | 0.605 |
| SII | 1035 (131– | 1098 (168–3743) | 766 (77–3337) | 0.370 |
| Albumin, g/dL | 3.4 (3.1–3.8) | 3.4 (3.1–3.9) | 3.3 (3.1–3.7) | 0.296 |
| Creatinine, mg/dL | 0.8 (0.6–0.9) | 0.8 (0.6–0.9) | 0.8 (0.6–1.0) | 0.868 |
| **Anthracycline exposure** |  |  |  |  |
| Doxorubicin-equivalent | 66 (49–92) | 69 (50–92) | 58 (49–89) | 0.110 |
| No. of recorded administration days | 2 (1–5) | 2 (1–6) | 1 (1–2) | <0.001 |
| More than one anthracycline agent | 31 (8.6) | 29 (9.5) | 2 (3.4) | 0.197 |
| Any liposomal doxorubicin | 17 (4.7) | 15 (4.9) | 2 (3.4) | >0.99 |
| Follow-up, days | 143 (86–244) | 138 (81–237) | 187 (124–309) | 0.003 |

Values are median (Q1–Q3) or n (%). P values were calculated using the Wilcoxon rank-sum test for continuous variables and the Fisher exact test for categorical variables; no adjustment was made for multiple comparisons. Malignancy categories were not mutually exclusive (15 patients had more than 1 malignancy). Insurance at the index admission is presented as a proxy for socioeconomic status because income is not recorded in MIMIC-IV. Medication use denotes inpatient medication orders active from 30 days before through 2 days after the index date. Follow-up duration was measured from the index date to the last follow-up echocardiogram within 365 days. Recorded doxorubicin-equivalent dose reflects inpatient exposure only and may underestimate cumulative anthracycline exposure. ACEI = angiotensin-converting enzyme inhibitor; ALC = absolute lymphocyte count; ARB = angiotensin receptor blocker; ARNI = angiotensin receptor– neprilysin inhibitor; COPD = chronic obstructive pulmonary disease; CTRCD = cancer therapy–related cardiac dysfunction; LMR = lymphocyte-to-monocyte ratio; LVEF = left ventricular ejection fraction; MIMIC-IV = Medical Information Mart for Intensive Care IV; NLR = neutrophil-to-lymphocyte ratio; PLR = platelet-to-lymphocyte ratio; RDW = red cell distribution width; SII = systemic immune-inflammation index.

### 3.2 Early Hematologic Response and Timing of CTRCD Detection

During the prespecified 0–30-day exposure window, lymphocyte percentage declined from a median of 10.9% to a nadir of 3.0%, corresponding to a median relative change of −73% (Q1–Q3: −89% to −44%). Crude relative changes were similar in patients with and without subsequent CTRCD (−71% [−88% to −25%] vs −73% [−89% to −48%]; P = 0.25). Relative change in absolute lymphocyte count was also similar between groups (P = 0.33) (Table 2). Descriptive paired trajectories of lymphocyte percentage and absolute lymphocyte count at baseline and day 30 are shown in Figures 2A and 2B.

**Table 2.** Early Hematologic Changes According to Subsequent CTRCD Status.

| Parameter | Overall<br>(n = 362) | No CTRCD<br>(n = 304) | CTRCD<br>(n = 58) | P<br>value |
| --- | --- | --- | --- | --- |
| **Lymphocyte percentage, %** |  |  |  |  |
| Baseline | 10.9 (5.4–22.6) | 11.6 (5.5–22.0) | 9.3 (5.0–24.0) | 0.436 |
| 0–30-day nadir | 3.0 (1.0–7.0) | 2.9 (1.0–6.6) | 4.0 (1.0–9.0) | 0.623 |
| Relative change, % | –73 (–89 to –44) | –73 (–89 to –48) | –71 (–88 to –25) | 0.246 |
| **Absolute lymphocyte count,**<br>**×10<sup>9</sup>/L** |  |  |  |  |
| Baseline | 0.74 (0.38–1.39) | 0.78 (0.39–1.37) | 0.60 (0.34–1.89) | 0.807 |
| 0–30-day nadir | 0.17 (0.08–0.32) | 0.19 (0.09–0.32) | 0.14 (0.08–0.20) | 0.046 |
| Relative change, % | –74 (–88 to –54) | –74 (–88 to –54) | –80 (–91 to –50) | 0.334 |
| **Other hematopoietic lineages,**<br>**relative change, %** |  |  |  |  |
| Neutrophil percentage | –75 (–93 to –38) | –75 (–93 to –36) | –76 (–94 to –50) | 0.515 |
| Monocyte percentage | –75 (–89 to –50) | –76 (–89 to –50) | –71 (–88 to –50) | 0.339 |
| White blood cell count | –91 (–97 to –72) | –90 (–97 to –70) | –95 (–98 to –82) | 0.017 |
| Platelet count | –69 (–88 to –46) | –68 (–87 to –44) | –79 (–91 to –56) | 0.037 |
| Immature granulocyte percentage | –33 (–55 to 0) | –31 (–50 to 0) | –47 (–69 to –17) | 0.017 |

Values are median (Q1–Q3). Relative change was calculated as $100 \times (\text{nadir} - \text{baseline})/\text{baseline}$ , where the nadir was the minimum value recorded during days 0–30. P values were calculated using the Wilcoxon rank-sum test comparing patients with and without subsequent CTRCD. CTRCD = cancer therapy–related cardiac dysfunction.

**Figure 2.**
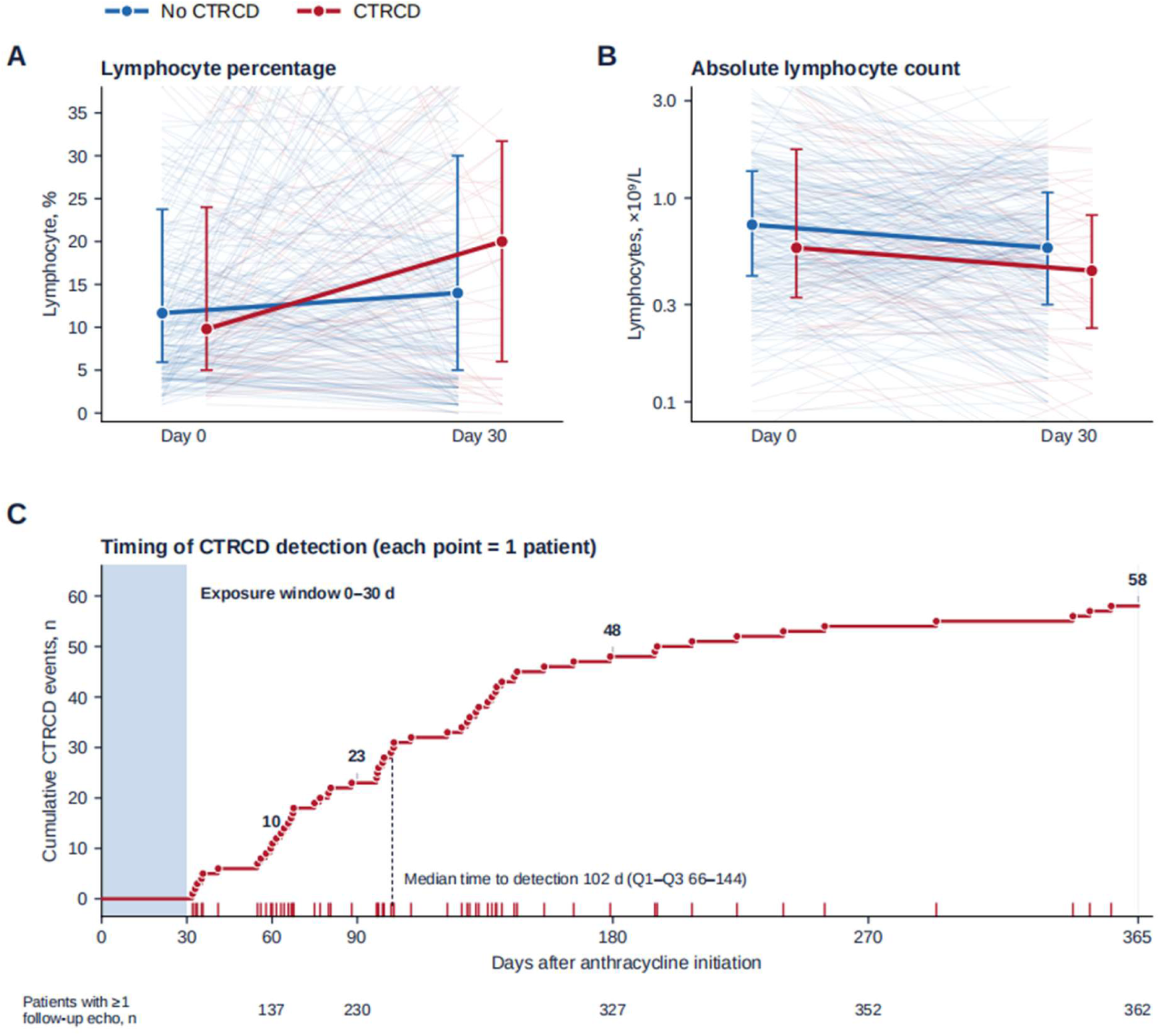
Early Lymphocyte Dynamics and CTRCD Timing. (A, B) Paired lymphocyte percentage and absolute lymphocyte count at baseline (day 0) and day 30, stratified by subsequent CTRCD status. Faint lines represent individual patient trajectories; points and error bars indicate medians and interquartile ranges. The absolute lymphocyte count is displayed on a logarithmic scale. These panels are descriptive and distinct from the primary exposure, defined as the relative change in lymphocyte percentage from baseline to the nadir during days 0–30. (C) Timing of CTRCD detection through day 365. Each point represents the first follow-up echocardiogram meeting the CTRCD definition in an individual patient (n = 58); red tick marks along the x-axis indicate individual event times. Shading indicates the 0–30-day exposure window, and outcome ascertainment began on day 31. Labels indicate cumulative CTRCD detections at 60, 90, 180, and 365 days. The dashed line indicates the median time to detection (102 days; interquartile range, 66–144 days). Numbers below the x-axis indicate the number of patients with at least one follow-up echocardiogram by the corresponding time point. CTRCD = cancer therapy–related cardiac dysfunction.

White blood cell count, platelet count, and immature granulocyte percentage showed nominal between-group differences in relative change, whereas neutrophil and monocyte percentage changes did not (Table 2). By design, outcome ascertainment began on day 31, after the 0–30-day exposure window. Cumulative detections reached 10 by 60 days, 23 by 90 days, 48 by 180 days, and 58 by 365 days (median time to detection, 102 days; Q1–Q3: 66–144) (Figure 2C).

### 3.3 Association of Early Lymphocyte Response With CTRCD

In the primary adjusted restricted cubic spline model, early lymphocyte percentage change was associated with subsequent CTRCD (overall P = 0.031; P for nonlinearity = 0.042). The adjusted odds of CTRCD were lowest near the median change and increased with a less pronounced decline. Compared with the median change (−73%), a less pronounced decline at the 90th percentile (−5%) was associated with higher odds of CTRCD (OR: 1.96; 95% CI: 1.12–3.42; P = 0.018) (Figure 3A; Supplemental Table S1).

**Figure 3.**
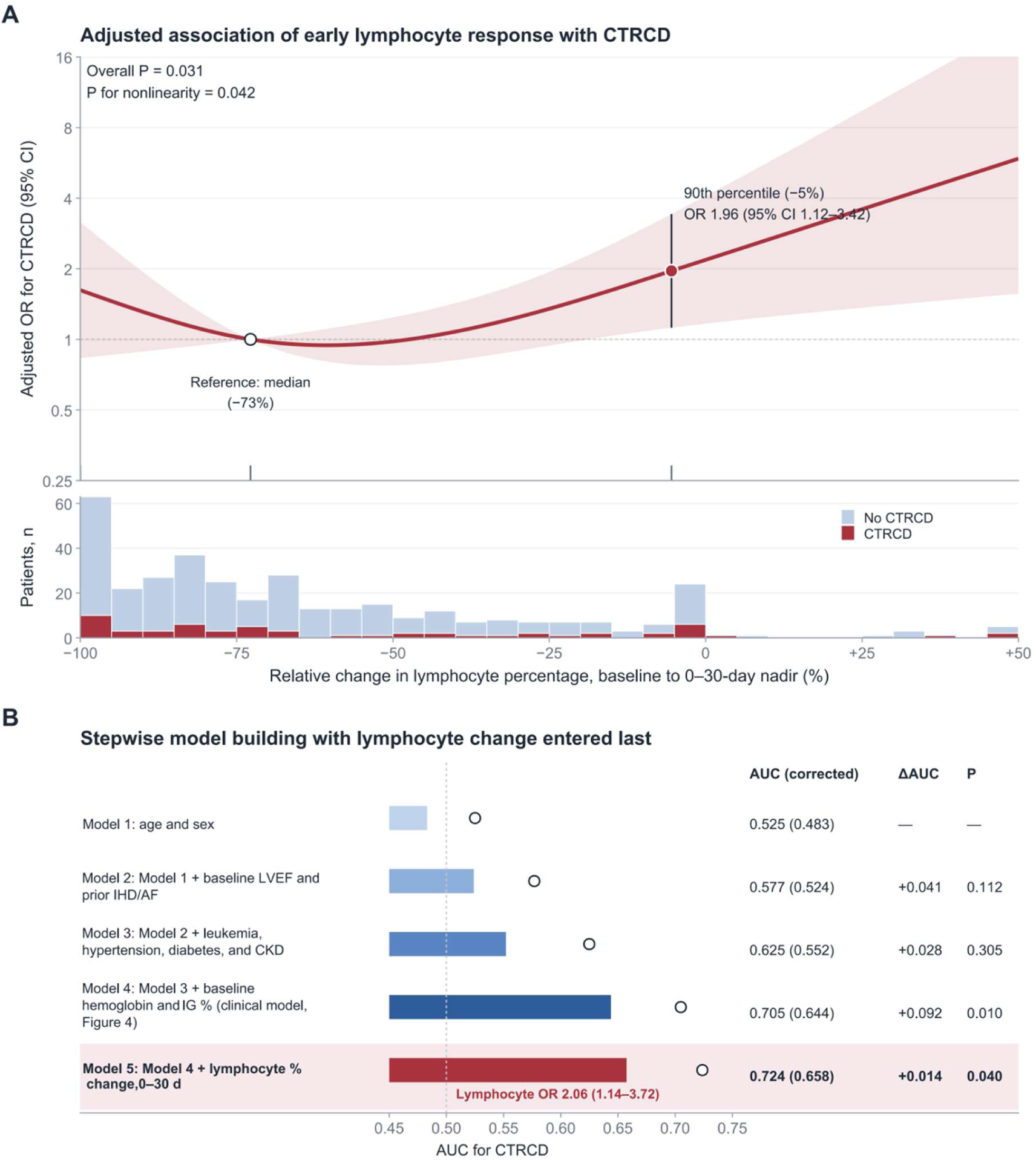
Association of Early Lymphocyte Response With CTRCD. A) Adjusted restricted cubic spline showing the association between relative change in lymphocyte percentage from baseline to the 0–30-day nadir and subsequent CTRCD. The median change (−73%) was the reference; the 90th-percentile change (−5%) was associated with an odds ratio (OR) of 1.96 (95% CI, 1.12–3.42). The overall association P value was 0.031, and the P value for nonlinearity was 0.042. Shading indicates the 95% CI, tick marks indicate knots at the 10th, 50th, and 90th percentiles, and the lower histogram shows the exposure distribution by CTRCD status. The model was adjusted for age, sex, baseline LVEF, prior ischemic heart disease or atrial fibrillation, leukemia, hypertension, diabetes mellitus, and chronic kidney disease. (B) Stepwise model building with early lymphocyte percentage change entered last. Each model includes the covariates in the preceding model; Models 4 and 5 correspond to the clinical and extended models, respectively, in Figure 4 and Table 3. Bars indicate optimism-corrected area under the receiver operating characteristic curve (AUC), and open circles indicate apparent AUCs. Predictions were averaged across 20 imputed datasets, with 250 bootstrap resamples per dataset. ΔAUC indicates the increase in optimism-corrected AUC from the preceding model; P values are pooled Wald tests of the terms added at each step. In Model 5, the OR comparing the 90th with the 50th percentile of relative lymphocyte percentage change was 2.06 (95% CI, 1.14–3.72). AF = atrial fibrillation; AUC = area under the receiver operating characteristic curve; CKD = chronic kidney disease; CTRCD = cancer therapy–related cardiac dysfunction; IHD = ischemic heart disease; LVEF = left ventricular ejection fraction; OR = odds ratio.

**Table 3.** Incremental Discrimination of Early Lymphocyte Change for CTRCD.

| Model | Apparent<br>AUC | Optimism-corrected<br>AUC | Change in<br>corrected<br>AUC | P for added<br>lymphocyte<br>terms |
| --- | --- | --- | --- | --- |
| Clinical model | 0.705 | 0.644 | — | — |
| Clinical model +<br>Lymphocyte % change (0–30<br>d) | 0.724 | 0.658 | +0.014 | 0.040 |

The clinical model included age, sex, baseline left ventricular ejection fraction (LVEF), prior ischemic heart disease or atrial fibrillation, leukemia, hypertension, diabetes mellitus, chronic kidney disease, baseline hemoglobin, and baseline immature granulocyte percentage. The extended model additionally included relative change in lymphocyte percentage from baseline to the 0–30-day nadir, modeled using a restricted cubic spline. Predicted probabilities were averaged across 20 multiply imputed datasets; baseline immature granulocyte percentage was missing in 110 patients. Optimism-corrected area under the receiver operating characteristic curve (AUC) values were estimated using 250 bootstrap resamples in each imputed dataset (5,000 resamples in total). The P value is a pooled Wald test of the 2 added lymphocyte spline terms, not a test of the difference in AUC. The continuous net reclassification improvement (NRI) was 0.36 (95% CI, 0.10–0.63), and the integrated discrimination improvement (IDI) was 0.018 (95% CI, –0.0002 to 0.037); 95% CIs for NRI and IDI were obtained using 500 bootstrap resamples. CTRCD = cancer therapy–related cardiac dysfunction.

The association was similar without covariate adjustment (OR: 1.95; 95% CI: 1.14–3.35) and after additional adjustment for baseline hemoglobin (OR: 1.99; 95% CI: 1.12–3.54), baseline immature granulocyte percentage (OR: 2.05; 95% CI: 1.15–3.64, with multiple imputation), or both (OR: 2.06; 95% CI: 1.14–3.72) (Supplemental Table S2), and it persisted after adjustment for changes in other hematopoietic lineages (ORs: 1.97–2.07) (Supplemental Table S3). Estimates were also similar after additional adjustment for glucocorticoid dose and granulocyte colony-stimulating factor (G-CSF) use during days 0–30 (OR: 2.05; 95% CI: 1.13– 3.73) and after inverse probability weighting for selection into the analysis cohort (OR: 1.92; 95% CI: 1.08–3.40). Adjustment for most additional covariates did not materially change the estimate (ORs: 1.86–2.30), whereas adjustment for recorded inpatient anthracycline administration days attenuated it (OR: 1.61; 95% CI: 0.91–2.84); the estimate adjusted for all additional covariates was 1.69 (95% CI: 0.91–3.14) (Supplemental Table S5; Supplemental Figure S1). No significant effect modification was observed across subgroups (all P for interaction ≥0.09), although subgroup estimates were imprecise (Supplemental Figure S2). Baseline absolute lymphocyte count, lymphocyte percentage, NLR, PLR, LMR, and SII were not associated with CTRCD (Supplemental Table S4).

### 3.4 Incremental Discrimination of Early Lymphocyte Response

Addition of early lymphocyte percentage change to the clinical model, which included baseline hemoglobin and immature granulocyte percentage, increased the apparent AUC from 0.705 to 0.724 and the optimism-corrected AUC from 0.644 to 0.658 (ΔAUC = 0.014; P = 0.040 for the lymphocyte spline terms), with a continuous NRI of 0.36 (95% CI: 0.10–0.63) and an IDI of 0.018 (95% CI: −0.0002 to 0.037) (Table 3; Figures 3B and 4). The bootstrap-corrected calibration slope of the extended model was 0.68, indicating some overfitting. In stepwise models, the optimism-corrected AUC was 0.483 with age and sex, 0.552 with all 8 clinical covariates, and 0.644 after addition of baseline hemoglobin and immature granulocyte percentage, before lymphocyte change was entered last (Figure 3B). When hemoglobin and immature granulocyte percentage were not included in the clinical model, lymphocyte change increased the optimism-corrected AUC from 0.552 to 0.587 (P = 0.031).

**Figure 4.**
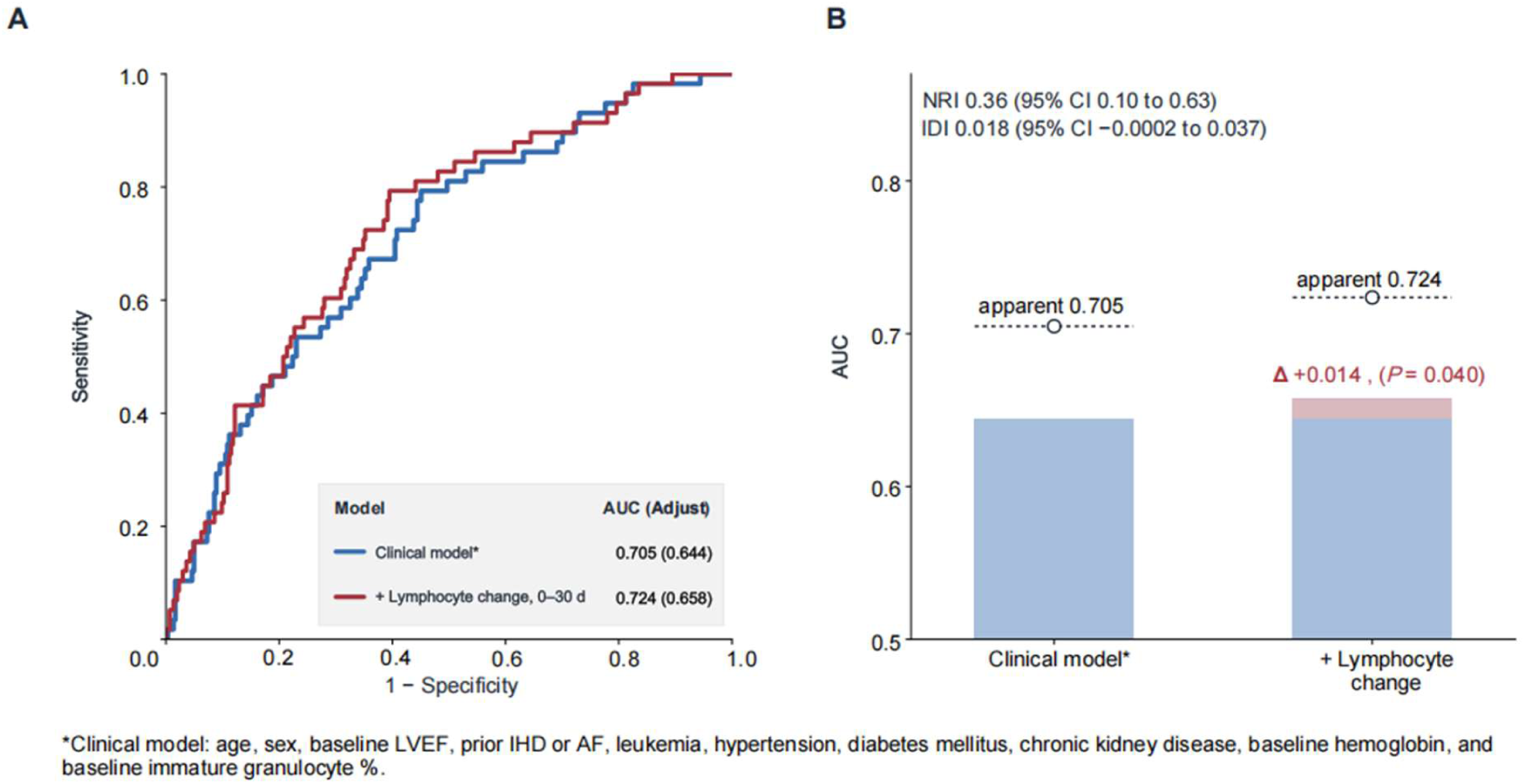
Incremental Discrimination of Early Lymphocyte Response. (A) Receiver operating characteristic curves for the clinical model and the model additionally including early lymphocyte percentage change modeled with a restricted cubic spline. The clinical model included age, sex, baseline LVEF, prior ischemic heart disease or atrial fibrillation, leukemia, hypertension, diabetes mellitus, chronic kidney disease, baseline hemoglobin, and baseline immature granulocyte percentage. Values indicate apparent AUCs, with optimism-corrected AUCs in parentheses. (B) Optimism-corrected AUCs (bars) and apparent AUCs (open circles). Adding early lymphocyte percentage change increased the apparent AUC from 0.705 to 0.724 and the optimism-corrected AUC from 0.644 to 0.658 (ΔAUC = 0.014). The P value of 0.040 refers to the pooled Wald test of the added lymphocyte spline terms and is not a direct test of the difference between AUCs. The continuous net reclassification improvement (NRI) was 0.36 (95% CI, 0.10–0.63), and the integrated discrimination improvement (IDI) was 0.018 (95% CI, −0.0002 to 0.037). AF = atrial fibrillation; AUC = area under the receiver operating characteristic curve; CTRCD = cancer therapy–related cardiac dysfunction; IDI = integrated discrimination improvement; LVEF = left ventricular ejection fraction; NRI = net reclassification improvement.

## 4 Discussion

In this retrospective cohort of anthracycline-treated patients, early within-patient change in lymphocyte percentage, rather than baseline lymphocyte status, was associated with subsequent CTRCD. A less pronounced decline during the first 30 days after anthracycline initiation was associated with approximately 2-fold higher odds of subsequent CTRCD. This association remained stable across sequential adjustment and after additional adjustment for changes in other hematopoietic lineages. Adding early lymphocyte change to clinical characteristics modestly improved optimism-corrected discrimination. Together, these findings suggest that the relevant lymphocyte signal may reside in the early treatment-emergent response rather than the pretreatment state.

Previous clinical studies have suggested that circulating inflammatory and hematologic markers may provide information about anthracycline-related cardiac injury, although the relevant markers and timing of assessment have varied across studies. A pilot cohort of women with HER2-positive breast cancer reported that higher pretreatment inflammatory indices, particularly the monocyte-to-lymphocyte ratio, were associated with subsequent CTRCD (6). During anthracycline exposure, Baruch et al. found that an elevated neutrophil-to-lymphocyte ratio was independently associated with subsequent deterioration in global longitudinal strain (10). Todorova et al. further showed that neutrophil-associated inflammatory proteins increased after the first cycle of doxorubicin-based chemotherapy and were higher in patients who developed cardiotoxicity (11). More recently, Gustafson et al. identified circulating endothelial activation and inflammatory biomarkers associated with subsequent CTRCD, with several vascular markers remaining elevated during treatment (12). Our previous study similarly found that post-treatment lymphocyte-based immunoinflammatory indices had greater discriminatory value for CTRCD than their pretreatment counterparts (7). Collectively, these observations suggest that the information carried by circulating inflammatory and hematologic markers may depend not only on the marker itself but also on the timing of assessment relative to treatment exposure.

Contemporary cardio-immunology evidence indicates that anthracycline-related cardiac injury involves heterogeneous innate and adaptive immune responses and that the biological effects of lymphocytes may depend on cellular subtype and activation state (5). Experimental work by Bayer et al. showed that doxorubicin increased circulating and myocardial IFNγ⁺CD8⁺T cells in mice, whereas CD8⁺T-cell depletion attenuated cardiac fibrosis and systolic dysfunction; increased circulating CD8⁺T cells were also observed after doxorubicin exposure in canine lymphoma and in a small group of patients with hematologic malignancies (13). Complementary work has implicated B cells, showing that B-cell depletion reduced doxorubicin-induced cardiac dysfunction, apoptosis, fibrosis, antibody deposition, and proinflammatory responses, whereas modulation of B-cell activation through the inhibitory receptor FcγRIIB conferred cardioprotection (14). These mechanisms highlight that the biological consequences of lymphocyte responses may depend on cellular subtype and activation state. Against this mechanistic background, our finding that a less pronounced early decline in lymphocyte percentage was associated with higher odds of subsequent CTRCD raises the possibility that the observed peripheral blood signal reflects treatment-emergent immune remodeling, potentially involving changes in the composition or activation state of specific lymphocyte populations.

Our findings also fit within the evolving view that cardiovascular risk during anthracycline therapy is dynamic rather than fixed at baseline. Contemporary reviews and cardio-oncology recommendations emphasize risk assessment across the treatment trajectory, integrating baseline cardiovascular vulnerability with surveillance as treatment exposure and patient status evolve (1–3,15–17). In a real-world cohort of 1,066 anthracycline-treated patients, Rivero-Santana et al. demonstrated that the baseline HFA-ICOS score effectively stratified the risk of clinically relevant anthracycline-related cardiovascular toxicity (18). More recently, the RE-ACT study showed that reassessment after anthracycline therapy using post-chemotherapy LVEF and cumulative anthracycline dose provided prognostic information complementary to baseline HFA-ICOS assessment, supporting a stage-specific approach to risk evaluation (4). Our findings extend this concept to an earlier treatment window by suggesting that a routinely available systemic hematologic response—captured by within-patient change in lymphocyte percentage during the first 30 days—may provide an additional layer of treatment-emergent information beyond conventional clinical characteristics.

The incremental discrimination observed in our study was modest, with the optimism-corrected AUC increasing from 0.644 to 0.658 when lymphocyte change was added to the clinical model (apparent AUC: 0.705 to 0.724). Accordingly, this finding should not be interpreted as establishing a clinical prediction model or as supporting the use of lymphocyte change as a stand-alone predictor. A systematic review of cardiotoxicity prediction models identified substantial methodological heterogeneity, limited external validation, and a high risk of bias in most published models, underscoring the need for rigorous independent validation before model-based risk estimates are adopted clinically (19). Experience with other treatment-emergent biomarkers also illustrates the distinction between detecting a biological response and achieving clinically useful discrimination. In the prospective Cardiac CARE study, serial high-sensitivity cardiac troponin I reflected treatment-related myocardial injury but was only weakly associated with subsequent LVEF change and showed limited predictive value for later ventricular dysfunction (20). In this context, the principal implication of our analysis is not that lymphocyte change constitutes a clinical prediction tool, but that early lymphocyte dynamics contain information not fully captured by conventional clinical characteristics.

## STUDY LIMITATIONS

This study has several limitations. Its retrospective, single-database design is subject to selection bias and residual confounding, and the predominance of hematologic malignancies may limit generalizability. Hematologic and echocardiographic assessments were clinically driven rather than protocolized, and incomplete availability of global longitudinal strain, cardiac biomarkers, and outpatient anthracycline exposure limited precise characterization of subclinical cardiac injury and cumulative treatment exposure. In addition, lymphocyte percentage is a compositional measure that cannot resolve immune-cell subsets or activation states, and concomitant therapies or treatment-related factors may have influenced lymphocyte kinetics. Although additional adjustment for glucocorticoid dose and G-CSF use did not change the estimate, adjustment for the number of recorded inpatient anthracycline administration days, which largely distinguished multiday infusional regimens, attenuated the association, suggesting partial confounding by treatment regimen that could not be fully resolved because outpatient treatment was not captured. Information on chest radiotherapy and concomitant HER2-targeted therapy was not reliably available. The E-value for the primary estimate was 2.15 (1.31 for the lower confidence limit), indicating that an unmeasured confounder would need to be associated with both early lymphocytes change and CTRCD by a risk ratio of at least 2.15 to fully explain the association. Finally, discrimination was modest and evaluated only internally; because hemoglobin and immature granulocyte percentage in the clinical model were identified in exploratory analyses, even the optimism-corrected AUCs may overestimate performance in new patients. We were unable to identify an independent public database that combined anthracycline administration records, serial blood counts, and quantitative LVEF for external validation, and independent prospective validation is required before clinical utility can be established.

## 5 Conclusion

A less pronounced early decline in lymphocyte percentage after anthracycline initiation was associated with higher odds of subsequent CTRCD and provided modest incremental information beyond clinical characteristics.

## Data Availability

The raw patient-level data used in this study are publicly available from the MIMIC-IV database hosted on PhysioNet ([https://physionet.org](https://physionet.org)). Access to MIMIC-IV requires completion of training and approval by the PhysioNet data use agreement. All analytical code generated for this study is available from the corresponding author upon reasonable request.

## Acknowledgments

ChatGPT was used for language editing. All content was reviewed by the authors.

## Non-standard Abbreviations and Acronyms

**AF** = atrial fibrillation; **ALC** = absolute lymphocyte count; **AUC** = area under the receiver operating characteristic curve; **CTRCD** = cancer therapy–related cardiac dysfunction; G-CSF = granulocyte colony-stimulating factor; IDI = integrated discrimination improvement; **IHD** = ischemic heart disease**; LMR** = lymphocyte-to-monocyte ratio; **LVEF** = left ventricular ejection fraction; **MIMIC-IV** = Medical Information Mart for Intensive Care IV; **NLR** = neutrophil-to-lymphocyte ratio; NRI = net reclassification improvement; PLR = platelet-to-lymphocyte ratio; **SII** = systemic immune-inflammation index

